# The subcortical morphology correlates of autistic traits in school-age children: a population-based neuroimaging study

**DOI:** 10.1101/2021.11.26.21266878

**Authors:** T.H. Sharp, M. Elsabbagh, A. Pickles, R. Bedford

**Affiliations:** King’s College London, Biostatistics and Health Informatics Department, Institute of Psychiatry, Psychology & Neuroscience, King’s College London, London, UK; Montreal Neurological Institute, Azrieli Centre for Autism Research, McGill University, Montreal, Canada; University of Bath, Department of Psychology, Bath, UK; King’s College London, Child and Adolescent Psychiatry Department, Institute of Psychiatry, Psychology & Neuroscience, King’s College London, London, UK

**Keywords:** Autism, autistic traits, subcortex, MRI, brain morphology, neuroimaging, ABCD

## Abstract

**Background:** There is emerging evidence that the neuroanatomy of autism forms a spectrum which extends into the general population. However, whilst several studies have identified cortical morphology correlates of autistic traits, it is not established whether morphological differences are present in the subcortical structures of the brain. Additionally, it is not clear to what extent previously reported structural associations may be confounded by co-occurring psychopathology. To address these questions, we utilised neuroimaging data from the Adolescent Brain Cognitive Development Study to assess whether a measure of autistic traits was associated with differences in child subcortical morphology, and if any observed differences persisted after adjustment for child internalising and externalising symptoms.

**Methods:** Our analyses included data from 7,005 children aged 9-10 years (female: 47.19%) participating in the Adolescent Brain Cognitive Development Study. Autistic traits were assessed using scores from the Social Responsiveness Scale. Volumes of subcortical regions-of-interest were derived from structural magnetic resonance imaging data.

**Results:** Overall, we did not find strong evidence for an association of autistic traits with differences in subcortical morphology in this sample of school-aged children. Whilst lower absolute volumes of the nucleus accumbens and putamen were associated with higher scores of autistic traits, these differences did not persist once a global measure of brain size was accounted for.

**Conclusions:** These findings from our well-powered study suggest that other metrics of brain morphology, such as cortical morphology or shape-based phenotypes, may be stronger candidates to prioritise when attempting to identify robust neuromarkers of autistic traits.

## Introduction

Autism is a neurodevelopmental condition characterised by difficulties in social interaction and communication, together with restricted interests and a tendency to engage in repetitive behaviours [1]. Aetiology is complex, with an interaction of genetic, environmental, and neurodevelopmental pathways thought to lead to clinical manifestation [2]. The first behavioural signs typically emerge in early childhood [3], and are accompanied by atypical development of brain structure, function, and connectivity, which are hypothesised to play a role in behaviours across the lifespan [4]. Characterising the neural correlates of autism has therefore remained a focus of the field [5].

Advances in neuroimaging technology in the last two decades have allowed the development of novel *in vivo* imaging methods of the human brain. Of these, structural magnetic resonance imaging (sMRI), has been most widely used to characterise the neuroanatomy of autism [6, 7]. The identification of a neural endophenotype could help inform clinical care, such as earlier diagnosis and intervention, and be used in the subtyping of individuals within the heterogenous autism umbrella [8]. In addition, a robust biomarker could help distinguish autism from other neurodevelopment disorders that have overlapping clinical features, enabling individuals to access targeted treatments. For example, previous work has demonstrated distinct subcortical and cortical group differences in individuals with autism in comparison to those with obsessive compulsive disorder (OCD) and attention-deficit/hyperactivity disorder (ADHD) [9]. Furthermore, if associations were identified to be causal, knowledge of the specific regions implicated could provide mechanistic insights and inform novel therapeutic strategies [10].

The neuroimaging literature demonstrates considerable heterogeneity regarding direction and effect size of brain morphology differences in autism [5, 11-13]. Whilst this may reflect the high level of aetiological and neurobiological heterogeneity across the autistic spectrum [14], methodological factors are thought to be a contributing factor. Firstly, existing studies tend to be in relatively small samples, leading to overestimation of effect sizes and low reproducibility [15]. Secondly, there is substantial heterogeneity in study design, including differences in participant characteristics such as age and symptom severity, covariates included, and neuroimaging outcomes assessed. Finally, analytic differences, such as variation in MRI acquisition, processing and analytic pipelines, impact results derived from individual studies [5]. Two mega-analyses (n∼3000) from the Enhancing Neuro-Imaging Genetics through Meta-Analysis (ENIGMA) consortium aimed to characterise the neuroanatomical correlates of autism, whilst addressing these issues [16]. Global measures were found to be higher in autistic participants, including intracranial volume (ICV), total grey matter, and mean cortical thickness. Regional differences in cortical thickness, including increases in the frontal regions and decreases in the temporal regions, were also observed. In contrast, no differences in cortical surface area were found [17]. Altered lateralised neurodevelopment was also revealed, with reduced regional asymmetry in cortical thickness and area, and increased asymmetry in the putamen [18].

An additional limitation of existing literature is the use of categorical diagnoses when assessing neural correlates of autism [6]. There has been little focus on identifying the brain morphology correlates associated with subclinical autism in typically developing populations. Subclinical autistic traits include social communication differences alongside restricted behaviour patterns, which do not cause difficulties for everyday functioning [19]. The behaviours associated with autism can therefore be considered continuous traits, which extend into the general population [20, 21]. Genetic studies have demonstrated that liability to autism influences typical variation in the population of social-emotional interaction and communication ability, providing further evidence for the importance of studying autism-related phenotypes in a quantitative manner [22]. Two longitudinal studies have examined cortical correlates of autistic traits in community-based samples, using measures from the Social Responsiveness Scale (SRS) [23]. Higher SRS scores were correlated with reduced regional cortical thickness including the right superior temporal sulcus [24] and the middle temporal gyri, ventral precentral and postcentral gyri, anterior cingulate and right frontopolar cortex [25], which remained stable from childhood to adolescence. In addition, a study within the Generation R cohort used vertex-wise modelling to demonstrate autistic traits were associated with properties of cortical morphology, including surface area, thickness, and gyrification [26]. Importantly, these differences persisted after exclusion of autism cases, providing further evidence for the extension of autistic traits into the general population. Whilst this work has begun to reveal the neurobiological differences associated with autistic traits, there remains a gap in the literature regarding differences in subcortical morphology. Given the robust reductions in regions including the pallidum, putamen, and nucleus accumbens reported in autism [17], and the plausible role of these structures in the socio-motivational, cognitive and motor symptoms seen in autism, it will be important to explore whether differences in these or other subcortical structures are observed in non-clinical samples.

In addition, children with autism are at increased risk of mental health issues and frequently present with problems in emotion, attention, and behaviour [27]. Whilst prevalence varies greatly, anxiety disorders, depression, OCD, ADHD, and specific phobias are most consistently reported as secondary psychiatric disorders co-occurring with autism [27-29]. Autistic traits have also been identified as a risk factor for poorer mental health, with associations appearing stronger in childhood than adulthood [30]. Whilst co-occurring psychopathology will confound behavioural-brain associations, such traits are not routinely controlled for in the existing autism neuroimaging literature [6]. The fact that the majority of studies are in clinical or community-based samples will bias towards a high occurrence of multiple diagnoses, and therefore there is a gap in the literature for the application of methods in epidemiological cohorts. The identification of brain morphology features that remain associated with autistic traits beyond correction for co-morbidities will help delineate the biological underpinning of autism from other neurodevelopment disorders with overlapping clinical features.

In our study we aim to expand on existing literature by exploring whether autistic traits are associated with differences in subcortical morphology, and whether any observed differences are explained by co-morbid psychopathology. We present the first population-based analysis of subcortical morphology associated with autistic traits in an epidemiological sample of 9-to-11-year-old children participating in the Adolescent Brain Cognitive Development^SM^ (ABCD) Study (n=7,005). Firstly, we explored whether a quantitative measure of autistic traits was associated with differences in child subcortical morphology. Secondly, to understand if any identified neural endophenotypes were specific to autistic traits, we tested whether associations persisted after controlling for co-occurring internalising and externalising symptoms.

## Methods

### Study Sample

The ABCD Study^®^ is a longitudinal study of brain development and child health. The study design and recruitment strategy have previously been described [31], but in brief, the study used school-based recruitment to enrol 11,875 children from 21 metropolitan sites across the United States. Children were aged between 9 and 11 years at time of enrolment, and they and their caregiver completed the baseline visit between the 1^st^ of October 2016 and the 31^st^ of October 2018, which consisted of questionnaires, clinical interviews, neurocognitive interviews, and a neuroimaging protocol. Exclusionary diagnoses include a current diagnosis of schizophrenia, a moderate/severe autism diagnosis, intellectual disability, or alcohol/substance use disorder.

This study received approval from the institutional review board of the University of Southern California. The ABCD Study obtained centralized institutional review board approval from the University of California, San Diego, and each of the 21 study sites obtained local institutional review board approval. Ethical regulations were followed during data collection and analysis. Parents or caregivers provided written informed consent, and children gave written assent. Data can be accessed through registration with the ABCD study at https://nda.nih.gov/abcd. The present analyses used data from the baseline (demographic information, co-occurring psychopathology) and follow-up phase one visits (SRS). A total of 11,878 children were recruited at baseline, and of these, 11,736 participated in sMRI scanning. As the ABCD cohort contains data from siblings, measures from a random sample of 7,875 unrelated individuals were used, of which 345 were excluded due to poor quality sMRI data. Of the remaining 7,521 participants, 7,005 had available data on autistic traits, and thus made up the present sample.

### Neuroimaging measures

Structural MRI scans were acquired at twenty-one sites across the United States using twenty-six different scanners from two vendors (Siemens and General Electric). Data were acquired when children were 10 years of age (range: 8.91 to 11 years). Methods and assessments were optimised and harmonised across ABCD study sites for 3-T scanners, which can be found detailed in a previous report [32]. In brief, T1-weighted structural scans with 1-mm isotropic resolution were collected using adult-size multi-channel coils, and harmonized image acquisition protocols for 3-Tesla Siemens, Phillips, and General Electric scanners at 21 sites. During the MRI scan children watched a child-friendly movie and scans were collected using real-time motion detection and correction. The quality control procedures were based on automated mean and SDs of brain values. In addition, trained raters inspected images for poor quality, artifacts such as motion-related ghosting, blurring, or ringing that prevent brain segmentation. Images were corrected for scanner-specific gradient distortions. Intensity inhomogeneity was corrected using a B1-bias field, and image intensity was harmonized across participants [32].

Structural MRI data were processed by the ABCD DAIRC (Data Analysis, Informatics & Resource Center) team using FreeSurfer v5.3 (http://surfer.nmr.mgh.harvard.edu/) and subjected to quality control procedures (detailed in Supplementary Methods S1) [33]. Post-processed FreeSurfer derived phenotypes from the ABCD cohort have been widely used in studies assessing predictors of interest with brain morphology outcomes, including those examining subcortical ROIs [34-37].

### Social Responsiveness Scale (SRS)

Autistic traits were assessed using the SRS, which is primarily used to assess the severity of social difficulties across the full range of severity in both autistic and non-autistic children [23]. Statistical properties of the SRS have previously been evaluated in a UK population-based sample of 5-to-8-year-old children [38]. In the ABCD sample, parents answered an 11-item abridged version of the questionnaire which has previously shown strong loadings on the first unrotated factor of a principal components analysis of the SRS in a paediatric sample [39]. Parents were asked to rate statements on a four-point Likert scale; 0 (not true); 1 (sometimes true); 2 (often true); and 3 (almost always true). It encompasses the three DSM-IV autism domains, with items relating to reciprocal social behaviour (e.g., “Has difficulty making friends, even when trying his or her best.”), stereotyped and repetitive behaviours (e.g., “Has more difficulty than other children with changes in his or her routine.”), and communication impairments (e.g., “Has trouble keeping up with the flow of normal conversation.”). Total raw summary scores from participants were calculated (mean 3.49 SD=0.49, range=0-39). As has been reported in other typically developing populations [40], scores were negatively skewed and not amenable to transformation.

### Covariates

Potential confounders of the exposure-outcome relationship were defined *a priori* based on previous literature. A minimum set of confounders required to adequately account for confounding were defined as: age, sex, ethnicity, cognition score, and a measure of socioeconomic status (family-level income). To assess the impact of co-occurring psychopathology, separate models were conducted with the inclusion of T-scores of externalising and internalising symptoms extracted from the Child Behaviour Checklist (CBCL) [41].

Demographic information (child sex, age at time of MRI, ethnicity, and total household income) were extracted from a demographics survey answered by the child’s main caregiver. Child cognitive ability was assessed using the NIH Toolbox® cognition measures (http://www.nihtoolbox.org) [42]. The toolbox consists of seven tasks that cover episodic memory, executive function, attention, working memory, processing speed, and language abilities and is used to generate a total cognitive score composite. The composite score demonstrates good test re-test reliability and validity in children [43].

The CBCL parent-report was used to measure internalising and externalising symptoms in participants [44]. This is a well-established parent-completed measure of emotional, behavioural and social problems in children and adolescents [45]. Composite scores of internalising and externalising problems were used for these analyses. Raw scores were converted to standardised t-scores, scaled so that fifty was average for child age and sex, with a standard deviation (SD) of ten points. Higher scores indicate increased behavioural and emotional problems.

### Statistical analysis

The association between SRS and brain morphology outcomes (volumes of the thalamus, caudate, putamen, pallidum, amygdala, hippocampus and NAcc) were modelled using seemingly-unrelated regression (SUR). The SUR system allows for a single model containing a number of linear equations, permitting correlation among the error terms.

Models were conducted in three steps to assess the impact of confounding variables. Model 1: adjustment for child age, sex, ethnicity, family income, and ABCD recruitment site. Model 2: model 1 with the addition of cognition score. Model 3: model 2 with the addition of externalising symptoms and internalising symptoms. Raw p-values were adjusted for multiple testing by using Holm correction. Sensitivity analyses explored whether differences in subcortical volume were explained by ICV. All analyses were conducted in Stata v16.0 [46], with the **-sureg** command utilised to conduct SUR. Correlation between individual ROIs were assessed using the **-pwcorr** command.

## Results

### Association of autistic traits with covariates

In our sample of children, male children of white ethnicity tended to score higher on the SRS compared to their peers (Table 1). A higher SRS score was negatively correlated with cognition score, and increased scores of total externalising and internalising problems. A strong negative gradient of family-level income with SRS score was observed. Children in the highest scoring SRS tertile demonstrated a lower ICV on average. The quality of sMRI data, which can be lower in autistic children due to increased participant motion inside the scanner [47], was not correlated with SRS score.

**Table 1:**
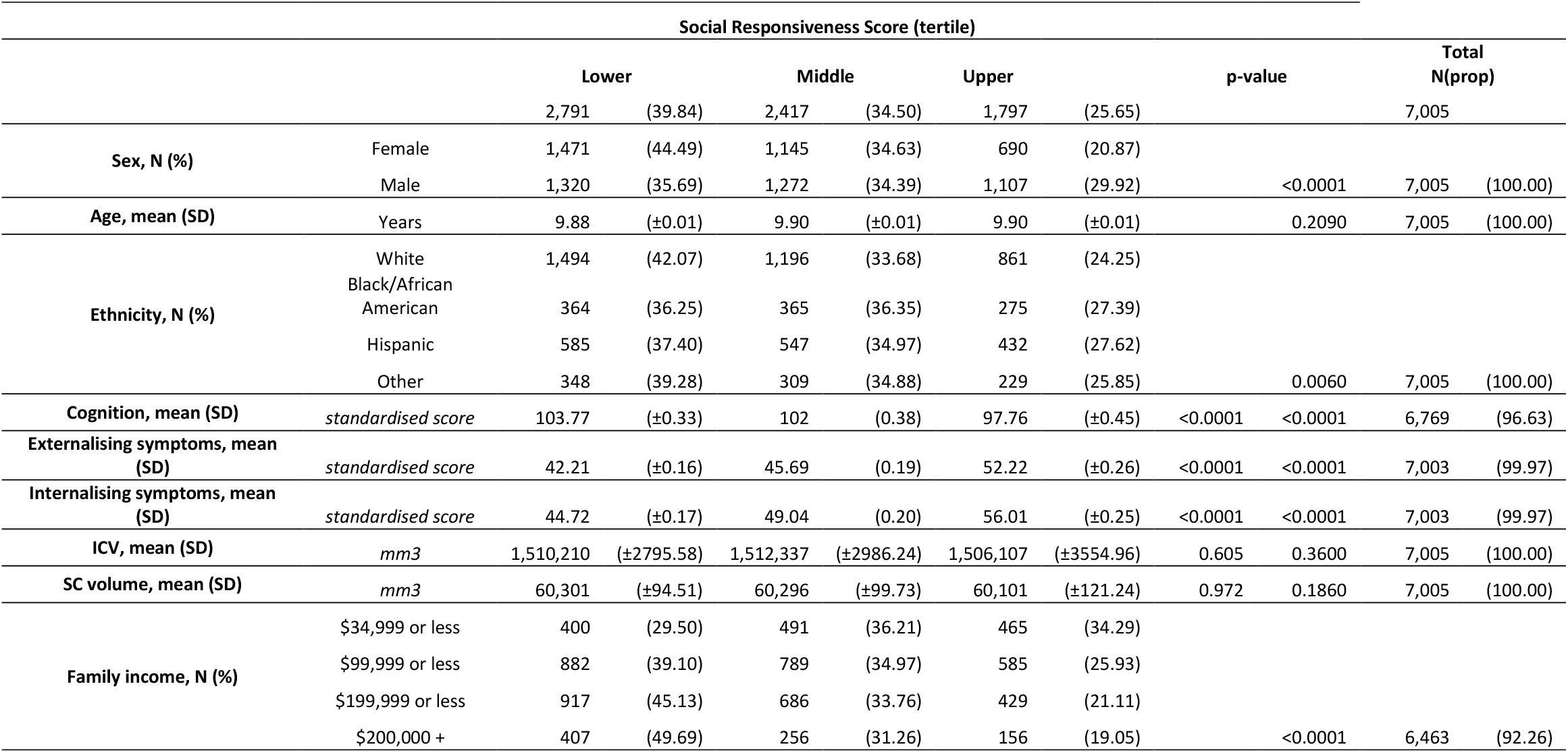
Distribution of covariates of interest stratified by SRS tertile. Reported p-values generated by univariate regression modelling for continuous variables and chi^2^ testing for categorical variables. ICV (intracranial volume), SC (subcortical).

### Association of global subcortical volume with covariates of interest

The association of covariates of interest with global subcortical volume were examined (Table 2). Male sex and older age were strongly predictive of higher subcortical volume. Non-white ethnicity, across all categories, was associated with lower subcortical volume. Children with increased scores on the total composite cognition score showed a higher subcortical volume. Whilst scores of total internalising problems showed little association, externalising symptoms were robustly associated with lower subcortical volume. Family-level income was positively associated with subcortical volume at all levels. Levels of correlation between individual subcortical ROIs was high across all comparisons justifying our use of SUR (Figure 1).

**Table 2:**
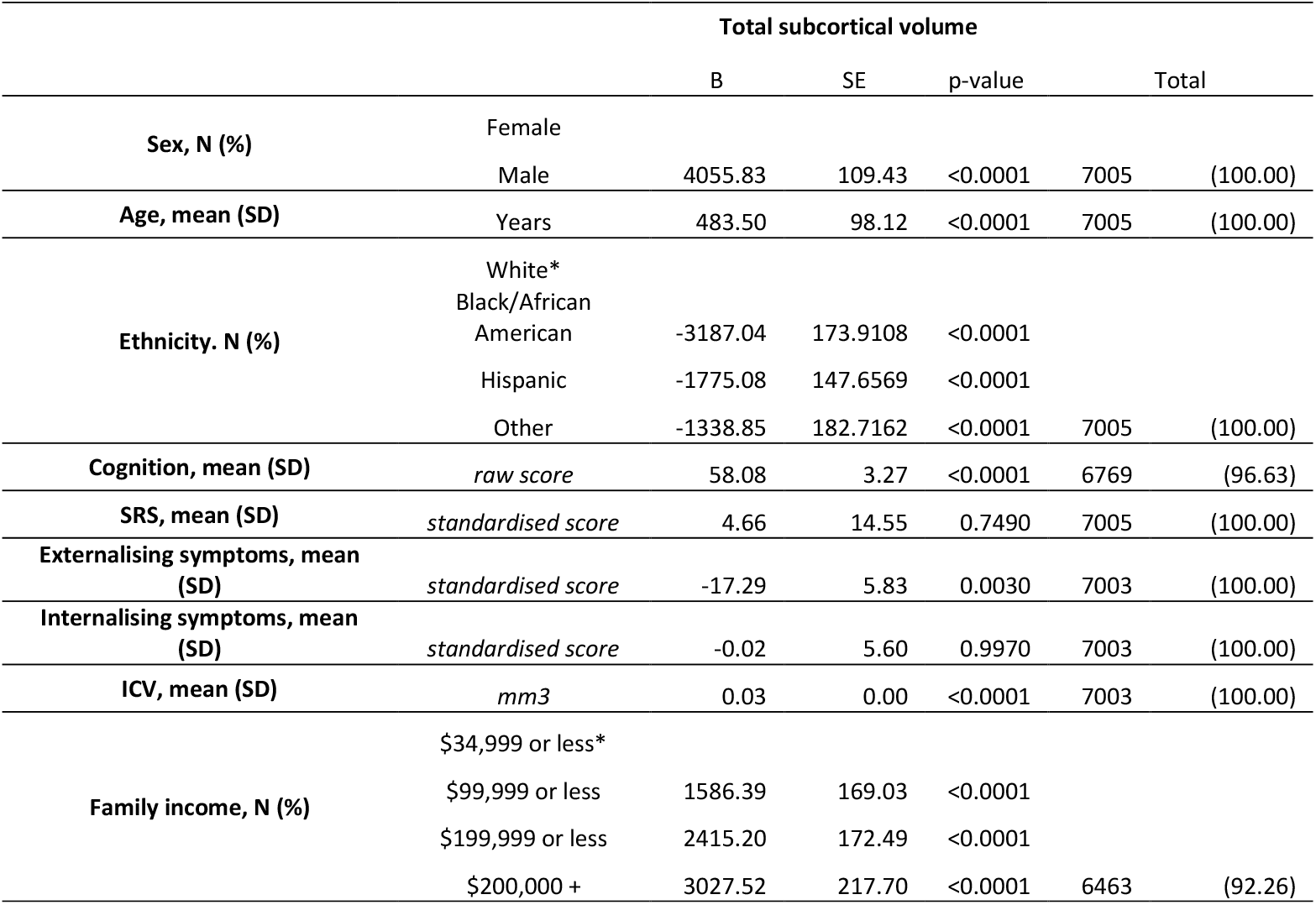
Results from univariate regression modelling of the association between covariates of interest and global neural outcomes. ICV (intracranial volume). *Indicator variable.

**Table 3:** Results from regression modelling of the association between the SRS and subcortical ROIs. Corrected p-values were generated using holm correction for multiple testing. NAcc: nucleus accumbens. Model 1 was adjusted for child age, sex, ethnicity, family income, and ABCD recruitment site. Model 2 was adjusted for the covariates included in model 1 with the addition of cognition score. Model 3 was adjusted for the covariates included in model 2 with the addition of externalising symptoms and internalising symptoms

|  | ROI | B | SE | p-value | holm p-value |
| --- | --- | --- | --- | --- | --- |
| Model 1 | thalamus | -7.87 | 3.82 | 0.039 | 0.118 |
|  | caudate | -7.60 | 3.12 | 0.015 | 0.059 |
|  | putamen | -9.38 | 3.67 | 0.011 | 0.053 |
|  | pallidum | -3.31 | 1.14 | 0.004 | 0.023 |
|  | amygdala | -1.14 | 1.09 | 0.294 | 0.294 |
|  | hippocampus | -4.42 | 2.29 | 0.053 | 0.118 |
|  | NAcc | -1.68 | 0.54 | 0.002 | 0.013 |
| Model 2 | thalamus | -3.77 | 3.85 | 0.327 | 0.674 |
|  | caudate | -5.74 | 3.16 | 0.069 | 0.336 |
|  | putamen | -6.78 | 3.70 | 0.067 | 0.336 |
|  | pallidum | -2.63 | 1.16 | 0.023 | 0.139 |
|  | amygdala | -0.51 | 1.10 | 0.644 | 0.674 |
|  | hippocampus | -2.81 | 2.31 | 0.225 | 0.674 |
|  | NAcc | -1.51 | 0.55 | 0.006 | 0.039 |
| Model 3 | thalamus | 0.53 | 4.33 | 0.903 | 1.000 |
|  | caudate | -4.88 | 3.55 | 0.169 | 1.000 |
|  | putamen | -5.20 | 4.17 | 0.213 | 1.000 |
|  | pallidum | -1.93 | 1.30 | 0.139 | 0.975 |
|  | amygdala | 0.95 | 1.24 | 0.441 | 1.000 |
|  | hippocampus | 1.40 | 2.60 | 0.590 | 1.000 |
|  | NAcc | -0.79 | 0.61 | 0.199 | 1.000 |

**Figure 1:**
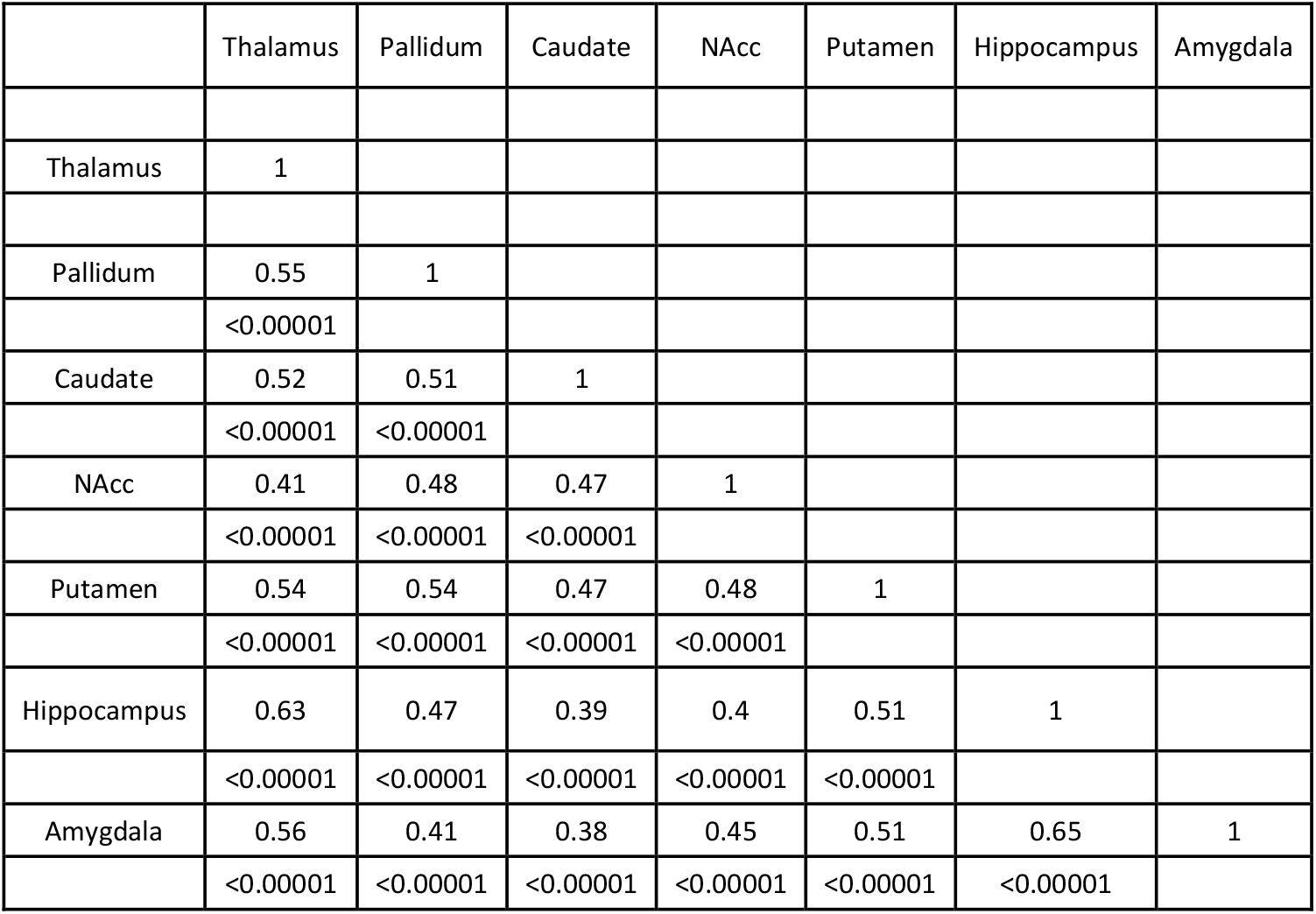
Correlation between subcortical regions-of-interest.

### Aim 1: Association of SRS traits with subcortical morphology

Overall, in our sample of 7,005 children from the ABCD study we found little evidence to suggest autistic traits are associated with disproportionate differences in volumes of seven subcortical structures (Figure 2).

**Figure 2:**
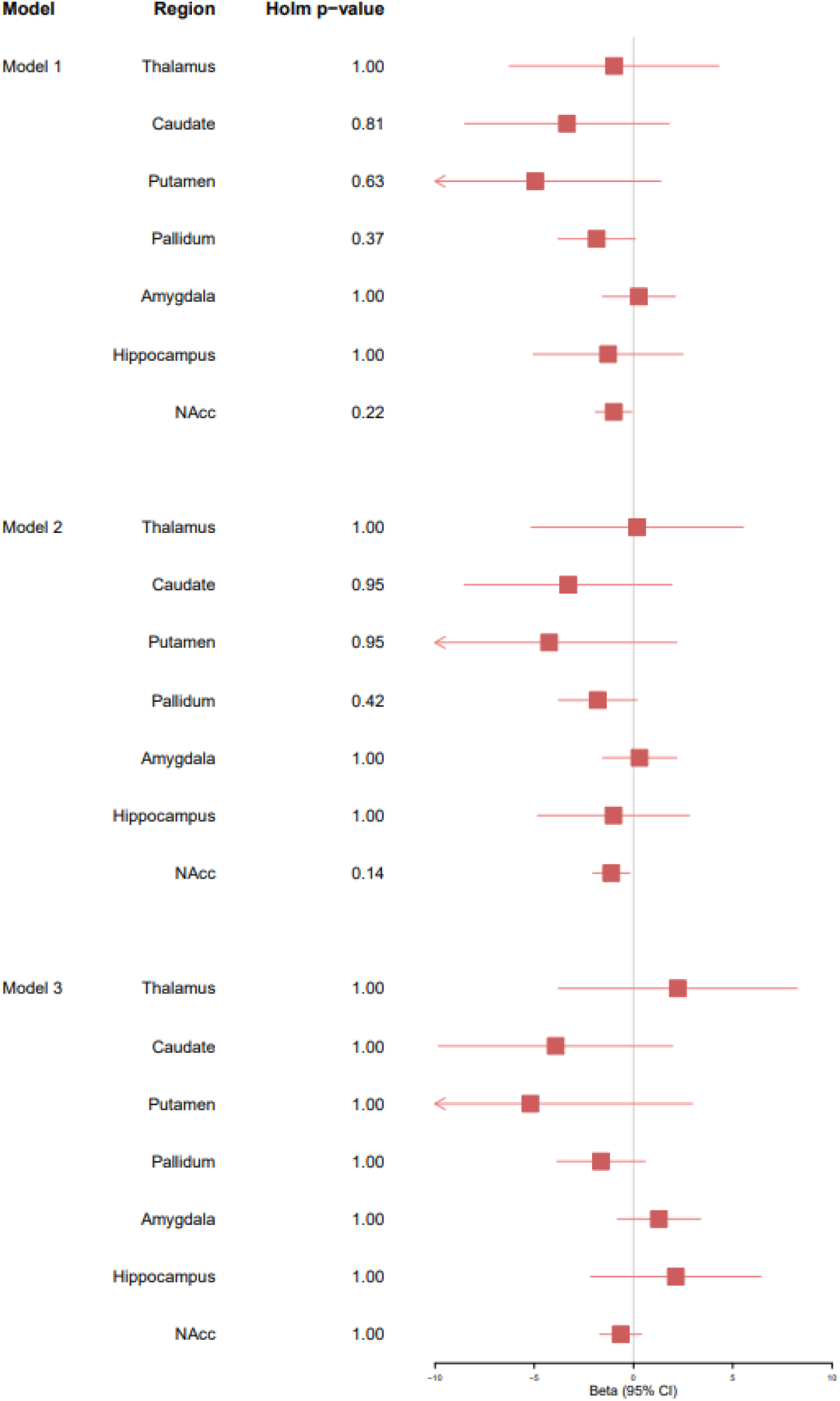
Forest plot depicting results from regression modelling of the association between the SRS and subcortical ROIs when adjusted for ICV. Corrected p-values were generated using holm correction for multiple testing. NAcc: nucleus accumbens. Model 1 was adjusted for child age, sex, ethnicity, family income, and ABCD recruitment site. Model 2 was adjusted for the covariates included in model 1 with the addition of cognition score. Model 3 was adjusted for the covariates included in model 2 with the addition of externalising symptoms and internalising symptoms

In model 1, adjusted for age, sex, ethnicity, ABCD recruitment site and family-level income, autistic traits were found to be predictive of absolute values of subcortical ROIs, with the strongest reductions with the NAcc (B= -1.68, SE=0.54, p_holm_=0.013) and pallidum (B= -3.31, SE=1.14, p_holm_=0.023). Suggestive associations were observed with the caudate and putamen (B= -7.60, SE=3.12, p_holm_=0.059 and B= -9.38, SE=3.67, p_holm_=0.053). The inclusion of total cognition score in model 2 attenuated these estimates towards the null, with only the reduction in NAcc volume remaining associated with SRS (β= -1.51, SE=0.55, p_holm_=0.039). In contrast, little difference in subcortical ROI volumes were observed once ICV was corrected for in sensitivity analyses (Table 4), suggesting these observed small differences are not beyond proportional differences of overall brain size in children.

**Table 4:** Results from regression modelling of the association between the SRS and subcortical ROIs with the inclusion of ICV. Corrected p-values were generated using holm correction for multiple testing. NAcc: nucleus accumbens. Model 1 was adjusted for child age, sex, ethnicity, family income, and ABCD recruitment site. Model 2 was adjusted for the covariates included in model 1 with the addition of cognition score. Model 3 was adjusted for the covariates included in model 2 with the addition of externalising symptoms and internalising symptoms.

|  | ROI | B | SE | p-value | holm p-value |
| --- | --- | --- | --- | --- | --- |
| Model 1 | thalamus | -0.98 | 2.69 | 0.72 | 1.00 |
|  | caudate | -3.35 | 2.63 | 0.20 | 0.81 |
|  | putamen | -4.95 | 3.23 | 0.13 | 0.63 |
|  | pallidum | -1.86 | 0.99 | 0.06 | 0.37 |
|  | amygdala | 0.27 | 0.94 | 0.78 | 1.00 |
|  | hippocampus | -1.28 | 1.92 | 0.51 | 1.00 |
|  | NAcc | -1.00 | 0.47 | 0.03 | 0.22 |
| Model 2 | thalamus | 0.18 | 2.73 | 0.95 | 1.00 |
|  | caudate | -3.29 | 2.67 | 0.22 | 0.95 |
|  | putamen | -4.25 | 3.27 | 0.19 | 0.95 |
|  | pallidum | -1.80 | 1.01 | 0.07 | 0.42 |
|  | amygdala | 0.30 | 0.95 | 0.75 | 1.00 |
|  | hippocampus | -1.01 | 1.95 | 0.61 | 1.00 |
|  | NAcc | -1.12 | 0.47 | 0.02 | 0.14 |
| Model 3 | thalamus | 2.35 | 3.07 | 0.44 | 1.00 |
|  | caudate | -3.75 | 3.01 | 0.21 | 1.00 |
|  | putamen | -4.03 | 3.68 | 0.27 | 1.00 |
|  | pallidum | -1.54 | 1.13 | 0.17 | 1.00 |
|  | amygdala | 1.33 | 1.07 | 0.21 | 1.00 |
|  | hippocampus | 2.23 | 2.19 | 0.31 | 1.00 |
|  | NAcc | -0.61 | 0.53 | 0.26 | 1.00 |

### Aim 2: Adjustment for co-morbid psychopathology

To assess whether any observed differences persisted after adjustment for co-occurring psychopathology, model 3 incorporated t-scores of total scores of externalising and internalising problems.

The observed weak reduction in absolute NAcc volume attenuated towards the null once externalising and internalising symptoms were accounted for (β= -0.79, SE=0.61, p_holm_=1.00), suggesting differences are not specific to autistic traits. Inclusion of externalising and internalising symptoms had little impact on effect estimates for the additional six subcortical ROIs. Similarly, sensitivity analyses examining the impact of inclusion of ICV did not alter effect estimates substantially.

## Discussion

There is emerging evidence that the neuroanatomy of autism falls along a continuum within the general population. Whilst several studies have assessed cortical phenotypes of autistic traits [25, 26], there remains a distinct gap in the literature regarding subcortical morphology. Thus, the primary aim of the present study was to investigate the association of autistic traits in childhood, measured by parent reported SRS score, with subcortical brain morphology. Our second aim was to test whether any observed differences were robust to adjustment for co-occurring psychopathology, measured as total scores of externalising and internalising symptoms. To our knowledge, this is the first such study to examine this association within the general population, and therefore represents a novel contribution to the current body of literature.

To summarise, in this study of school aged children in the ABCD cohort, we did not find strong evidence for an association of autistic traits with differences in the subcortical volumes assessed, with results compatible with the null hypothesis and generally wide confidence intervals throughout. Whilst we observed lower absolute volumes of the NAcc and putamen in those scoring higher on the SRS, this attenuated towards the null once overall brain size was accounted for. As univariate analyses had demonstrated children in the upper tertile of SRS scores had on average a lower ICV, this suggests the observed differences were not beyond that of proportional differences in brain size of the children in our sample. This finding of a reduced global measure of brain volume is in line with other studies assessing the neural correlates of autistic traits [26, 48].

In the ABCD sample, being male or of white ethnicity was associated with higher SRS scores. It has been previously reported that in samples from the general population, male children tend to have higher SRS scores than female [38, 48, 49]. In contrast, there is little published literature regarding distribution across ethnic groups, and therefore this is an area which requires further investigation.

The existing literature is composed predominantly of studies using a case-cohort design. Most notably, findings from the ENGIMA consortium identified lower volumes of the pallidum, putamen, and NAcc in autistic participants compared to controls [9]. *Post-hoc analyses* demonstrated these differences were related to the degree of autism symptom severity, measured by scores extracted from the Autism Diagnostic Observation Schedule (ADOS) [50]. Although it must be noted that there are qualitative differences between the SRS and ADOS [51], and that these findings from the ENGIMA consortium have not yet been replicated, we had hypothesised we may see similar effects of a smaller magnitude focussed on these specific ROIs when examining the correlates of SRS scores in our sample. One possible explanation for our null results are that the differences in subcortical morphology observed in autism cases may represent neurobiology associated with a higher degree of autistic symptoms that meet the criteria for a clinical diagnosis.

A further study from the Generation R neuroimaging cohort, whilst predominantly focussed on cortical morphology, examined one subcortical ROI in relation to autistic traits [26]. The authors utilised a sample from 9- to 12-year-olds in the Netherlands (n=2400), examining amygdala volume in relation to SRS scores. In line with our findings, amygdala volume was found to not differ significantly with SRS score when covariates were accounted for. In contrast, strong evidence was found for differences in metrics of cortical morphology, including lower gyrification, thickness and surface area, suggesting autistic traits in this sample are primarily associated with cortical, rather than subcortical ROI, differences.

Our second aim was to explore the role of co-occurring psychopathology, to understand if neural phenotypes were specific to autistic traits or simply a reflection of generalised psychopathology. Inclusion of these covariates had little impact on effect estimates, however, given that we found little association with SRS scores alone and that univariate analyses did not demonstrate strong associations of these covariates with our outcomes of interest, this is unsurprising.

It is important to note that whilst we did not detect significant group differences in subcortical ROIs, it is possible these volumetric measures are not sensitive to what may be more subtle differences exerted by autistic traits in the general population. Aggregate measures such as volume do not fully capture the complexity of subcortical structures and may be insensitive to specific local effects, or obscure heterogenous local effects by averaging out subtle differences in shape [52]. This is particularly true for phenotypes which are likely characterised by specific associations with functionally distinct subfields of subcortical structures, such as traits of autism. Therefore, our lack of detectable volumetric differences in subcortical ROIs may be due to analytic methods, which do not allow for these subtler differences to be assessed. Whilst no studies have specifically used shape-based methods when assessing the subcortical correlates of autism, it has been demonstrated that for other neurobehavioral phenotypes, these methods provide more information than volumetric methods alone. For example, a recent study examining the subcortical alterations associated with major depressive disorder found little difference in subcortical volumes, beyond that of lower hippocampal volume [53]. In contrast, subsequent analyses using shaped-based methods identified specific effects localised to regions of the amygdala and hippocampus associated with patients in comparison to controls [54]. Complementary analyses, using shape-based analytic methods, will therefore be necessary to understand if autistic traits are associated with more sensitive markers of difference in subcortical morphology.

When interpreting our findings, several limitations must be noted. Firstly, our analyses were based on sMRI data obtained at a single time point, limiting our analyses to a cross-sectional design. Autism correlates with altered neurodevelopmental trajectories, which has specifically been demonstrated for subcortical morphology [55]. It will therefore be important to continue this work where repeated neuroimaging measures are available, to understand if autistic traits are associated with individual or group differences in trajectories of subcortical volumes. As the ABCD cohort is an ongoing, longitudinal study, it will provide the ideal sample to continue examining these trends as further data is released [56]. Secondly, neuroimaging measures and SRS scores were not contemporaneous, however, given the relatively short time period between clinics, and that autistic traits have been showed to remain stable over time [40, 57], this will likely have had limited impact. Thirdly, as the ABCD cohort excluded participants with a moderate or severe autism diagnosis (based on whether a child’s caregiver reported they did not attend mainstream school), the average severity of autistic traits will be artificially lower than in the general population, and therefore findings may be biased towards the null. Fourthly, as information regarding whether children had received a clinical diagnosis of autism was not available, it was not possible to conduct sensitivity analyses excluding these participants. Finally, it is important to note that the SRS is contaminated by general behavioural problems [51], and therefore may not be wholly indicative of autism-specific symptoms.

These limitations must be contrasted against the multiple strengths of our study. Firstly, data were drawn from a large population-based cohort with autistic traits measured continuously. The use of a dimensional approach, rather than a case-cohort design, is better suited to the idea of an autism spectrum, and allowed us to test whether the underlying subcortical neurobiology of these traits extends into the general population. In addition, the ABCD cohort is socioeconomically, ethnically, and racially diverse, whilst being relatively homogenous regarding the age of participants. This allowed the generation of a representative estimate of the association of autistic traits with subcortical morphology, minimising the selection bias that has hindered previous studies in clinical samples. In addition, the wealth of phenotypic data available allowed us to control for all identified potential confounders of the exposure-outcome relationship, a significant source of bias in existing studies. In addition, utilising data from the ABCD cohort allowed a large sample size, with a total of 7,005 included participants, two-fold greater than that of the largest published study in this area. In conclusion, in our population-based sample of 9- to 11-year-olds, we did not find evidence for an association between autistic traits and subcortical volumetric differences. Our findings suggest that other metrics of brain morphology may be better targets when attempting to identify robust biomarkers of these traits and potential targets for intervention. However, it will be important to replicate these findings in an independent cohort, in addition to conducting complementary shape-based analysis of subcortical structures, as these methods may be more sensitive to subtle differences associated with autistic traits in childhood.

## Data Availability

All data produced in the present work are contained in the manuscript.

## Acknowledgements

Data used in the preparation of this article were obtained from the Adolescent Brain Cognitive Development^SM^ (ABCD) Study (https://abcdstudy.org), held in the NIMH Data Archive (NDA). This is a multisite, longitudinal study designed to recruit more than 10,000 children age 9-10 and follow them over 10 years into early adulthood. The ABCD Study® is supported by the National Institutes of Health and additional federal partners under award numbers U01DA041048, U01DA050989, U01DA051016, U01DA041022, U01DA051018, U01DA051037, U01DA050987, U01DA041174, U01DA041106, U01DA041117, U01DA041028, U01DA041134, U01DA050988, U01DA051039, U01DA041156, U01DA041025, U01DA041120, U01DA051038, U01DA041148, U01DA041093, U01DA041089, U24DA041123, U24DA041147. A full list of supporters is available at https://abcdstudy.org/federal-partners.html. A listing of participating sites and a complete listing of the study investigators can be found at https://abcdstudy.org/consortium_members/. ABCD consortium investigators designed and implemented the study and/or provided data but did not necessarily participate in the analysis or writing of this report. This manuscript reflects the views of the authors and may not reflect the opinions or views of the NIH or ABCD consortium investigators.

## Financial Statement

ME is funded by the Canadian Institutes of Health Research and Fonds de Recherche du Québec. RB was supported by a King’s Prize Fellowship (204823/Z/16/Z). AP was part funded by the National Institute for Health Research (NIHR) Maudsley Biomedical Research Centre at South London and Maudsley NHS Foundation Trust and King’s College London and SI award NF-SI-0617-10120. The views expressed are those of the author(s) and not necessarily those of the NHS, the NIHR or the Department of Health and Social Care.

## Supplementary Material

### Supplementary Methods

FreeSurfer extracts cortical and subcortical region-of-interests (ROIs) based on the Desikan-Killiany atlas [58]. The automated pipeline consists of co-registration based on a template reference surface, motion correction and averaging. Any intensity variation across the image due to magnetic field heterogeneity is corrected, and the skull stripped from the normalised intensity image. Images are then segmented using a connected components algorithm, where connectivity is not permitted across established cutting planes. Any holes within white matter are filled, producing a single volume for each hemisphere. Surfaces are then covered with a tessellation formed of polygons and smoothed to minimise distortion to form a surface-based reconstructed model of the cortical sheet. The grey/white matter, and white matter/pial surface boundaries are then delineated on derived images. Cortical thickness measures are defined as the average distance between the grey-white matter boundary and the pial surface. Surface measures are defined as the total area of tessellation classified within each boundary. Volumetric segmentation are used to delineate and label global (total brain volume, subcortical volume) and regional (nucleus accumbens, amygdala, caudate nucleus, hippocampus, pallidum, putamen, and thalamus) measures [33].

The resulting output was then visually examined by a trained DAIC technician, who rated them from zero to three in five categories: motion, intensity homogeneity, white matter underestimation, pial overestimation, and magnetic susceptibility artifact. From this an overall “pass” or “fail” score was generated. Participants whose images failed QC were excluded from the present analyses [58].

